# Sympathetic Innervation Modulates Ventricular Repolarization and Arrhythmia Vulnerability After Myocardial Infarction

**DOI:** 10.64898/2026.04.07.26350356

**Authors:** Javier Villar-Valero, Lledó Nebot, David Soto-Iglesias, Giulio Falasconi, Antonio Berruezo, Bastiaan J.D. Boukens, Beatriz Trenor, Juan F. Gomez

## Abstract

**Background:** Sympathetic modulation via the stellate ganglia is increasingly recognized as a contributor to ventricular arrhythmogenesis after myocardial infarction. However, the mechanisms by which autonomic remodeling interacts with chronic infarct substrates to shape arrhythmic vulnerability remain incompletely understood.

**Objectives:** To test the hypothesis that left- and right-sided stellate ganglion–mediated SNS modulation differentially reshapes ventricular arrhythmic vulnerability in chronic post-infarcted substrates, and that the RVI detects changes in vulnerability beyond conventional stimulation-based inducibility.

**Methods:** Fourteen patient-specific ventricular models with chronic post-infarcted remodeling were reconstructed from imaging data. A total of 336 simulations were performed under different combinations of stellate ganglion modulation, border zone remodeling, and fibroblast density. Arrhythmic vulnerability was quantified using 3D RVI mapping during paced rhythms and compared with conventional stimulation-based inducibility outcomes.

**Results:** Stellate ganglion modulation induced marked, regionally heterogeneous changes in repolarization timing, resulting in lower and more negative RVI values in vulnerable regions. More negative RVI values reflect increased propensity for wavefront–waveback interaction and reentry initiation. Across the cohort, stellate modulation consistently decreased RVI_min_, even when inducibility outcomes remained unchanged. These findings indicate that SNS modulation can create a substrate more permissive to reentry independently of whether ventricular arrhythmia is triggered during programmed stimulation.

**Conclusions:** Stellate ganglion–mediated sympathetic modulation dynamically reshapes ventricular arrhythmic vulnerability in chronic post-infarcted substrates. RVI provides a spatially resolved, vulnerability-based metric that complements inducibility testing by revealing autonomic–substrate interactions underlying arrhythmogenesis

**Condensed Abstract:** Sympathetic modulation via the stellate ganglia can alter ventricular repolarization and promote arrhythmogenesis after myocardial infarction, yet clinical responses remain heterogeneous. Using 14 patient-specific post-infarction ventricular models, we simulated left- and right-sided stellate modulation across combinations of border zone remodeling and fibrosis (336 simulations). Stellate modulation induced regionally heterogeneous repolarization shortening and reduced RVI values, even when programmed stimulation inducibility remained unchanged. These findings suggest that RVI captures substrate-level vulnerability beyond binary induction testing and may improve mechanistic assessment of autonomic–substrate interactions in chronic infarct substrates.

## 1. Introduction

Ventricular Arrhythmias (VAs) after Myocardial Infarction (MI) remain a leading cause of sudden cardiac death, despite advances in revascularization, pharmacological therapy, and device implantation [1]. While the arrhythmogenic role of infarct scar and peri-infarct tissue is well established, increasing evidence suggests that the Sympathetic Nervous System (SNS) plays a critical modulatory role in arrhythmia initiation and maintenance [2]. In this context, Cardiac Sympathetic Denervation (CSD) targeting the stellate ganglia has emerged as a therapeutic option for patients with refractory VAs [3]. However, clinical outcomes remain highly heterogeneous, with some patients experiencing substantial benefit and others showing limited or no response.

Traditional approaches to studying post-MI arrhythmia risk often rely on stimulation protocols to assess inducibility [4]. However, inducibility is a protocol-dependent outcome with only two possible results: either Ventricular Tachycardia (VT) is induced, or VT is not induced, which may fail to capture more subtle, yet clinically relevant, changes in Electrophysiology (EP) vulnerability. Some substrates are intrinsically arrhythmogenic and sustain reentry under most conditions, whereas others remain non-inducible despite significant EP modulation. In this context, metrics that directly quantify local susceptibility to reentry, independently of a specific stimulation protocol, may provide a more sensitive and mechanistic assessment of arrhythmogenic risk.

In this study, we investigate how sympathetic modulation, mediated by the left and right stellate ganglia, alters ventricular arrhythmogenic vulnerability, as quantified by the Reentrant Vulnerability Index (RVI). Using personalized post-MI ventricular models, we assess changes in RVI induced by stellate-related remodeling across different scar locations and Border Zone (BZ) conditions. We further examine how these RVI changes relate to arrhythmia induction when a complete stimulation protocol is applied. By focusing on stellate-induced modulation of the arrhythmogenic substrate rather than inducibility alone, this work positions RVI as a mechanistic and predictive marker of sympathetic-driven arrhythmic risk after MI.

#### WHAT IS KNOWN?

- Sympathetic modulation via the stellate ganglia plays a key role in ventricular arrhythmogenesis, particularly in post-infarction substrates.
- Arrhythmic risk is traditionally assessed using inducibility testing, which provides a binary and protocol-dependent measure of vulnerability.
- Repolarization heterogeneity is a recognized mechanism underlying reentry initiation in structurally remodeled myocardium.

#### WHAT THE STUDY ADDS

- Patient-specific computational models demonstrate that stellate ganglion modulation induces spatially heterogeneous APD shortening, amplifying repolarization gradients at scar-border interfaces.
- The reentrant vulnerability index (RVI) reveals dynamic changes in arrhythmic substrate that are not captured by conventional inducibility testing.
- The interaction between infarct geometry and regional sympathetic modulation determines whether autonomic input promotes or suppresses arrhythmic risk.
- These findings provide a mechanistic framework to interpret the variable clinical response to autonomic therapies such as stellate ganglion blockade or cardiac sympathetic denervation.

## 2. Methods

This study builds upon a set of patient-specific ventricular models previously developed and validated in [4]. Here, we extend that framework to investigate how sympathetic modulation mediated by the stellate ganglia alters arrhythmogenic vulnerability, with a specific focus on changes in the RVI.

### 2.1. Patient-specific ventricular models and tissue characterization

Anatomical reconstructions, mesh generation, fiber orientation assignment, and baseline EP properties were obtained from the cohort described in Villar-Valero et al. (2025) [4]. Briefly, ventricular geometries were reconstructed from LGE-MRI, including a segmental investigation of the effects of sympathetic modulation on the dense scar core and infarct BZ, following a clinically established 5SD thresholding strategy [5]. Fiber orientation was assigned using an atlas-based approach derived from Streeter et al. [6].

Fourteen post-infarction cases with distinct scar distributions and localizations were selected for the present study. These models provide a consistent and previously validated anatomical and EP substrate upon which to investigate the effects of sympathetic modulation.

### 2.2. Representation of stellate ganglion modulation

Regions under the influence of the left stellate ganglion (LSG) and right stellate ganglion (RSG) were explicitly defined for each patient. The left ventricle was subdivided according to the standard 17+11 AHA segmentation scheme [7], and each segment was assigned to LSG or RSG innervation based on established clinical knowledge [8], with LSG predominance over anterior–lateral regions and RSG predominance over septal–inferior regions [3]. These assignments were reviewed in collaboration with EP specialists. A complete list of segments assigned to LSG and RSG territories is provided in Supplementary Figure S1 to ensure reproducibility.

Figure 1 provides an overview of the 14 patient-specific ventricular models included in this study, highlighting the spatial relationships between the infarct scar and regions associated with left- and right-sided stellate ganglion innervation. This visualization was used not only for qualitative inspection, but also to classify each case according to the anatomical overlap between scar tissue and sympathetic territories. Specifically, patients were grouped into three categories: (i) cases with scars overlapping both LSG- and RSG-innervated regions, (ii) cases with scars predominantly located within LSG territories, and (iii) cases with scars predominantly located within RSG territories. This classification allowed subsequent analyses to relate changes in reentrant vulnerability, quantified by the RVI, to the relative distribution of scar and sympathetic modulation rather than to inducibility outcomes alone.

**Figure 1:**
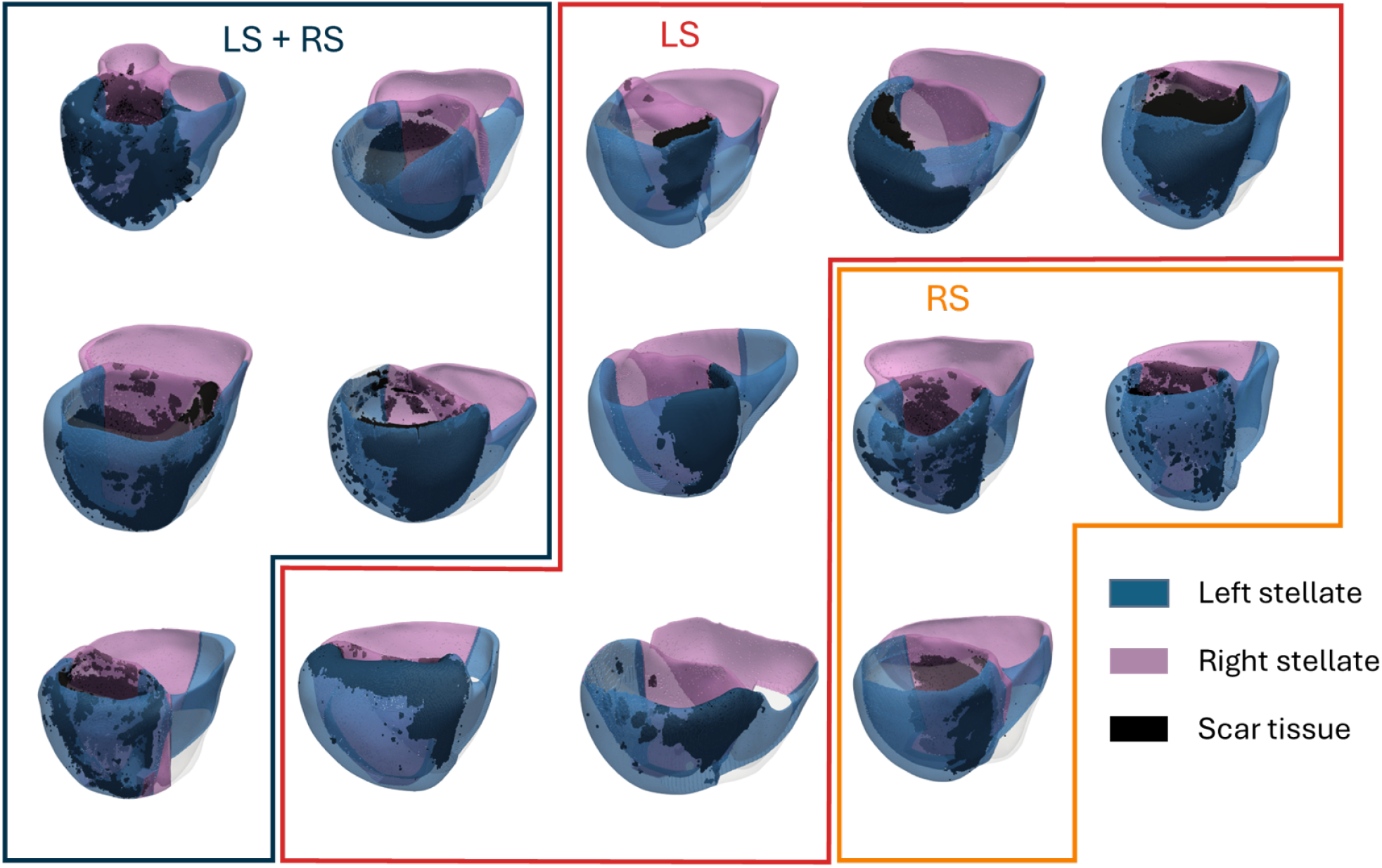
Reconstruction of the 14 biventricular models used in this study, illustrating the spatial distribution of regions associated with the left stellate ganglion (LSG, blue), the right stellate ganglion (RSG, pink), and infarct scar tissue (black). Cases are additionally grouped according to scar location: the blue box includes patients with scars overlapping both LSG- and RSG-innervated regions, the red box includes cases with scars predominantly located within LSG territories, and the orange box includes cases with scars predominantly located within RSG territories.

Sympathetic modulation was modelled phenomenologically as a regional shortening of action potential duration (APD) within the ventricular territories affected by stellate ganglion modulation. Rather than explicitly modelling the full intracellular *β*-adrenergic signalling cascade, this approach was designed to isolate the electrophysiological consequences of APD shortening and its spatial heterogeneity in the presence of complex, patient-specific post-infarction substrates.

APD shortening was implemented by scaling the conductance of the slow delayed rectifier potassium current (*I*_*Ks*_) in the O’Hara–Rudy human ventricular myocyte model [9]. The scaling factor was calibrated to achieve an average APD reduction of approximately 30%, representing a robust modulation of ventricular repolarization. Although this magnitude of APD shortening has been reported under conditions of acute ischemia with sympathetic activation [10], it was adopted here to probe an upper-bound effect of repolarization shortening in chronically remodelled ventricles. This phenomenological representation enables a controlled investigation of how spatially heterogeneous APD shortening interacts with scar geometry, border zone remodeling, conduction velocity heterogeneity, and fibrosis to shape ventricular arrhythmic vulnerability at the organ scale.

### 2.3. Electrophysiological Modeling

Electrical propagation was simulated using a monodomain formulation solved with the ELVIRA finite-element framework [11]. Cellular EP was represented using the modified O’Hara–Rudy human ventricular model [9, 12]. Dense scar tissue was treated as non-excitable by setting the conductivity to zero.

Post-infarction remodeling in the BZ followed the formulation described by Villar-Valero et al [4], which included ionic current alterations leading to a 17% APD prolongation representative of post-infarction remodeling [13]. To reproduce reduced intercellular coupling, the diffusion coefficients were reduced by 50% in both the longitudinal and transverse directions. Importantly, this reduction was calibrated to account for spatial resolution–dependent numerical effects, ensuring physiologically realistic conduction slowing without introducing discretization-related artifacts, as reported [14].

Diffuse fibrosis was incorporated using the MacCannell fibroblast model [15]. Different fibrosis burdens (0%, 30%, 50%, and 80%) were explored by adjusting fibroblast density and reducing effective diffusion by 50%, reflecting progressive electrical uncoupling in fibrotic tissue.

### 2.4. Simulation Design and Study Configurations

Each ventricular model was simulated under multiple configurations that combined three sympathetic states (no stellate modulation, LSG modulation, and RSG modulation), the presence or absence of BZ ionic remodeling, and four levels of fibrosis density. This resulted in 24 configurations per patient and 336 simulations in total.

This design allowed isolation of the individual and combined effects of sympathetic modulation, structural remodeling, and tissue heterogeneity on arrhythmia vulnerability.

### 2.5. Reentry Vulnerability Index

Arrhythmia vulnerability was primarily quantified using the RVI, computed following the methodology described by Campos et al. [16]. The RVI was evaluated prior to the application of any premature stimulation in order to characterize the baseline electrophysiological vulnerability of the substrate independently of inducibility protocols.

The concept underlying the RVI was originally introduced by Coronel et al., who referred to it as the fibrillation factor, linking spatial dispersion of repolarization to arrhythmogenesis and susceptibility to reentrant activity [17]. The metric was later translated into clinical practice by the group of Taggart et al., who coined the term RVI and demonstrated its ability to identify localized regions with high reentry susceptibility in patients [18].

Activation time (AT) was defined as the time at which the transmembrane potential crossed -20 mV during the upstroke. Repolarization time (RT) was computed as AT plus APD_90_ at each node. In this framework, the RVI at each node was defined as the difference between the local RT and the ATs of neighboring nodes within an 8 mm radius, consistent with prior clinical and computational implementations of RVI [19]. More negative RVI values indicate an increased likelihood of wavefront–waveback interaction, reflecting a higher propensity for reentry initiation. Three-dimensional RVI maps were generated for all simulated configurations, enabling a quantitative and spatial assessment of how stellate ganglion modulation alters the arrhythmogenic electrophysiological substrate.

In addition to spatial RVI mapping, three quantitative metrics were derived for each configuration: (i) the minimum RVI value (RVI_min_), (ii) the percentage of myocardial nodes with RVI<0, and (iii) the 1st percentile of the RVI distribution. These metrics were used to quantify global shifts in arrhythmic vulnerability independently of inducibility outcomes.

### 2.6. Arrhythmia Induction Protocol

To relate RVI changes to functional arrhythmia manifestation, an S1–S2 stimulation protocol previously described in Villar-Valero et al [4] was applied. Sustained reentry was defined as activation persisting for at least three complete cycles.

Importantly, arrhythmia inducibility was used as a secondary validation endpoint, allowing assessment of whether stellate-induced changes in RVI translated into increased or decreased susceptibility to reentry when appropriately triggered.

## 3. Results

### 3.1. Effect of stellate ganglion modulation on repolarization heterogeneity

Figure 2 illustrates the impact of stellate ganglion modulation on ventricular repolarization in two representative patients. Panel A shows repolarization time (RT) maps for each patient under the three sympathetic configurations (NoSG, LSG, and RSG). Compared to the baseline condition, activation of either stellate ganglion produced marked, regionally localized shortening of RTs, leading to increased spatial heterogeneity across the ventricular myocardium.

**Figure 2:**
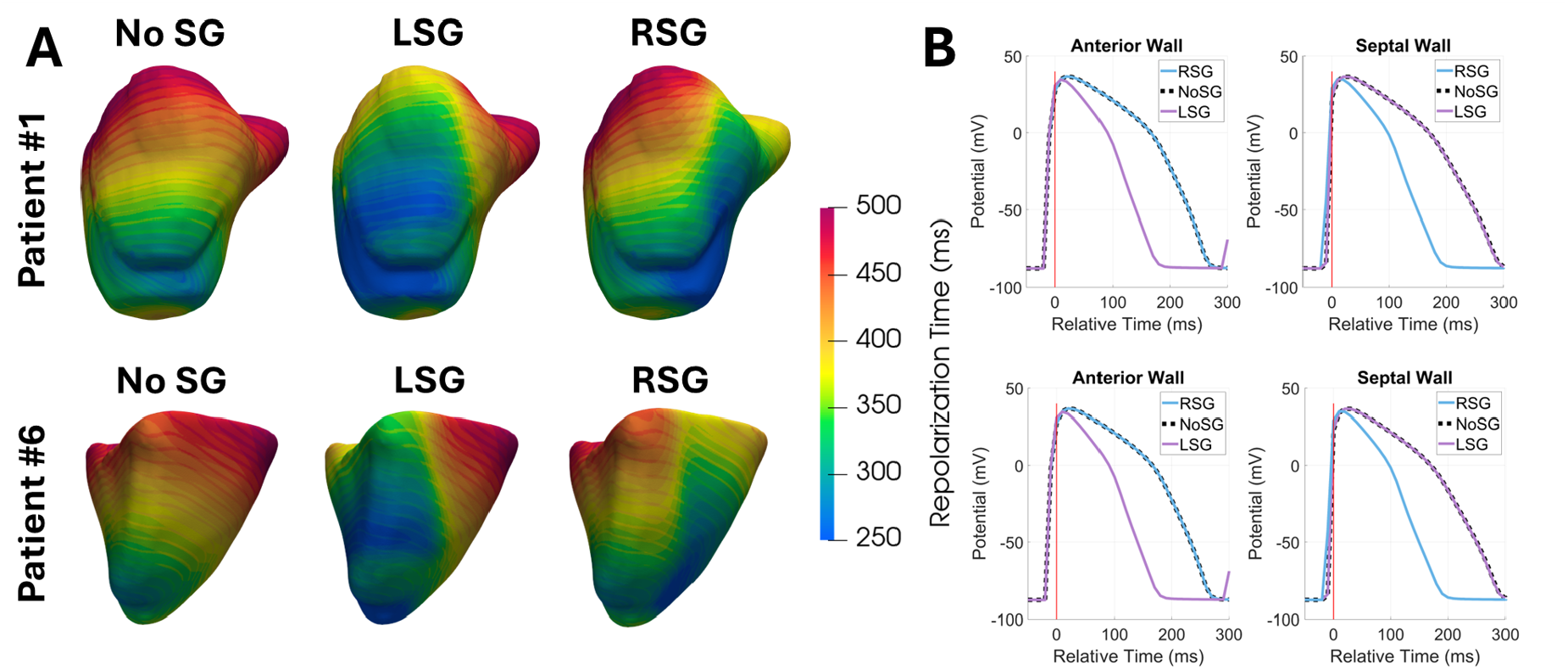
Effect of stellate ganglion modulation on ventricular repolarization and action potential duration. Panel A: Repolarization time (RT) maps for two representative patients (top row: Patient #1; bottom row: Patient #6) under three conditions: no stellate modulation (NoSG), left stellate ganglion (LSG) activation, and right stellate ganglion (RSG) activation. RTs are referenced to the last baseline pacing stimulus preceding the event. Panel B: Action potential traces extracted from two representative nodes (anterior wall and septal wall). Traces were aligned at AT to facilitate direct comparison of action potential duration.

To further illustrate the underlying cellular mechanism, Panel B displays action potential traces from two representative nodes: one in the anterior wall and one in the septal wall. Action potentials were temporally aligned at AT to allow direct comparison of APD across configurations. In each comparison, the baseline NoSG condition (black dotted traces) is consistently superimposed on the trace from the wall region unaffected by stellate modulation, serving as a reference. As expected, stellate ganglion modulation induced a pronounced APD shortening only in the node located within the corresponding innervated territory (anterior wall for LSG, septal wall for RSG). In contrast, when stellate modulation was not applied (NoSG) or when the applied modulation did not affect the selected node, APD remained unchanged and overlapped with the baseline trace.

These differences were not restricted to the infarct BZ but extended to remote healthy regions innervated by the corresponding stellate ganglion. As a consequence, sharper repolarization gradients emerged at the interface between healthy myocardium and slowly conducting BZ tissue. This stellate-driven increase in repolarization heterogeneity constitutes the EP substrate underlying the subsequent changes observed in reentrant vulnerability, as quantified by the RVI.

### 3.2. Global arrhythmia outcomes and mechanistic classification

Across the fourteen post-infarction models, a total of 336 simulations were performed under different combinations of stellate modulation, BZ remodeling, and fibroblast density. A comprehensive summary of inducibility outcomes for all configurations is provided (see Supplementary Material Table S1).

Table 1 presents a condensed patient-level overview relating the anatomical location of the infarct to the predominant mechanism associated with VA occurrence. While in some patients arrhythmia induction was primarily determined by scar structure or BZ remodeling, in others a clear dependence on sympathetic modulation mediated by the stellate ganglia was observed.

**Table 1:** Patient-level summary of infarct location and dominant mechanism associated with VA inducibility. Infarct location is reported using simplified anatomical left ventricular regions.

| ID | Infarct location | In silico VA | Clinical VA | Dominant mechanism |
| --- | --- | --- | --- | --- |
| P01 | Global LV | ✓ | ✓ | Scar structure |
| P02 | Inferior/Lateral | ✓ | ✗ | Stellate modulation |
| P03 | Inferior/Lateral | ✓ | ✓ | Stellate modulation |
| P04 | Apical | ✓ | ✓ | Scar structure |
| P05 | Anterolateral + Inferior/Lateral | ✓ | ✗ | Scar structure |
| P06 | Anterolateral + Inferior/Lateral | ✗ | ✗ | Scar structure |
| P07 | Inferior/Lateral + Anterolateral | ✓ | ✓ | Border zone |
| P08 | Inferior/Lateral | ✓ | ✗ | Scar structure |
| P09 | Inferior/Lateral | ✗ | ✓ | Scar structure |
| P10 | Apical | ✓ | ✓ | Border zone |
| P11 | Inferior/Lateral | ✓ | ✓ | Stellate modulation |
| P12 | Anterolateral | ✓ | ✓ | Border zone |
| P13 | Global LV | ✓ | ✓ | Stellate modulation |
| P14 | Inferior/Lateral | ✓ | ✓ | Border zone |

In patients classified as stellate-dominant, VA inducibility was facilitated by sympathetic activation acting on a remodeled substrate. Although the specific laterality of stellate involvement may differ across infarct locations, this simplified classification emphasizes the dominant electrophysiological driver of arrhythmogenesis rather than the spatial relationship between neural input and scar.

Importantly, this categorical classification highlights the heterogeneity of mechanisms across patients but does not fully capture differences in arrhythmic vulnerability within a given inducibility outcome. This limitation motivates the use of continuous vulnerability metrics, such as the RVI, to characterize pro-arrhythmic electrophysiological substrates beyond binary inducibility.

### 3.3. Stellate-driven modulation of reentrant vulnerability: representative cases

Figure 3 illustrates the effect of stellate ganglion modulation on reentry vulnerability in two representative patients with distinct mechanisms.

**Figure 3:**
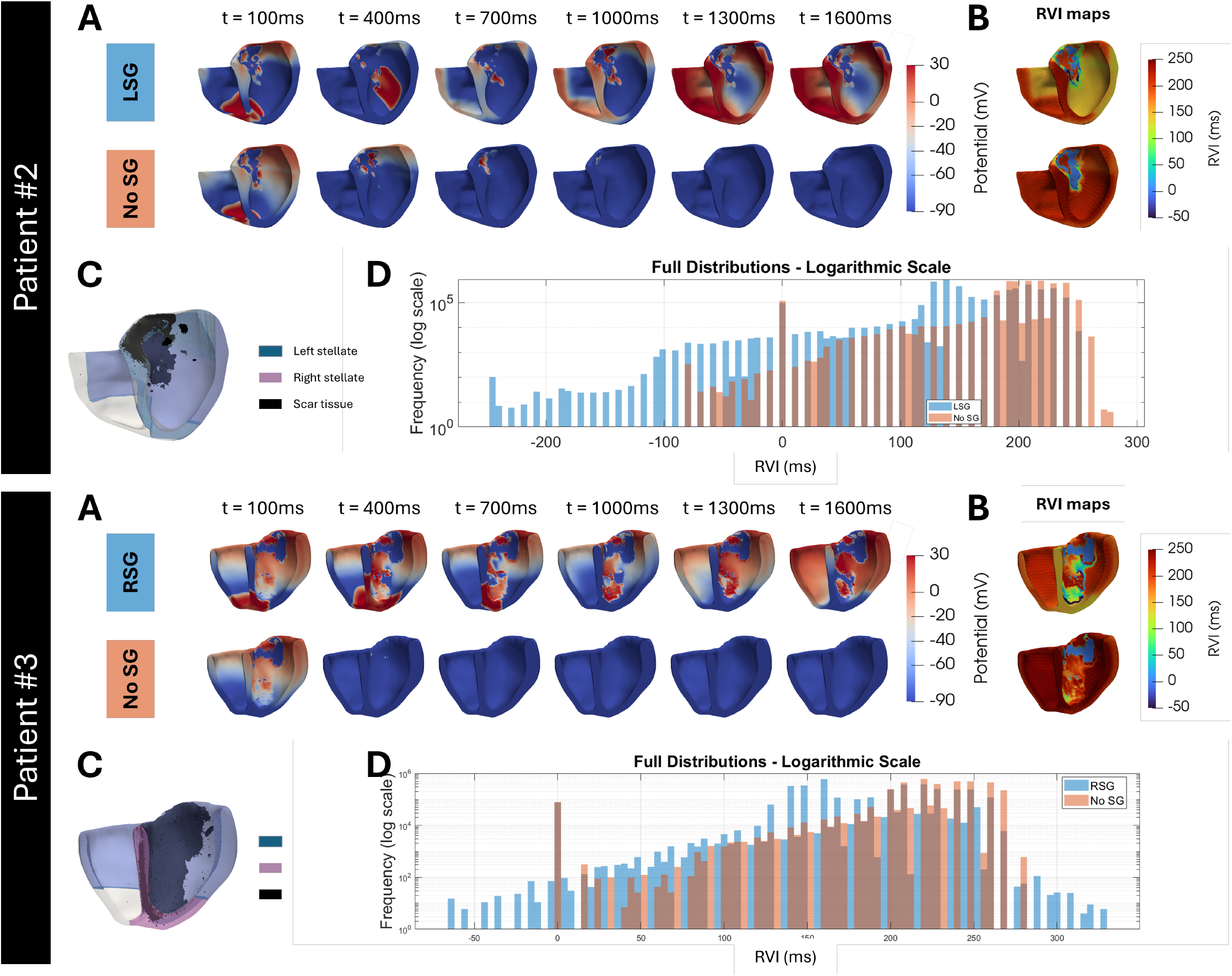
Representative cases illustrating distinct mechanisms by which stellate ganglion modulation alters reentrant vulnerability. For each patient, Panel A shows activation snapshots during the stimulation protocol (top: inducible condition; bottom: non-inducible condition). Panel B presents the corresponding RVI maps, highlighting regions of low and negative RVI associated with reentry initiation. Panel C displays the patient-specific ventricular anatomy, including infarct scar (black) and regions innervated by the left (blue) and right (pink) stellate ganglia. Panel D shows histograms of the RVI distributions across the entire mesh for conditions with and without stellate modulation (logarithmic scale).

In Patient #2, whose infarct scar was located within the left stellate territory, activation of the LSG led to a pronounced increase in repolarization heterogeneity near the BZ. Shortened action potentials in healthy LSG-innervated regions were juxtaposed with delayed activations within slow-conducting BZ channels, resulting in strongly negative RVI values. This shift was accompanied by an apparent leftward displacement of the global RVI distribution, a substantial increase in negative values, and culminated in reentry initiation during the stimulation protocol.

To gain mechanistic insight into how stellate modulation translates into changes in reentry vulnerability, we analyzed local activation dynamics in Patient #3. Representative wavefront propagation maps and simulated action potential traces are shown in Figure 4.

**Figure 4:**
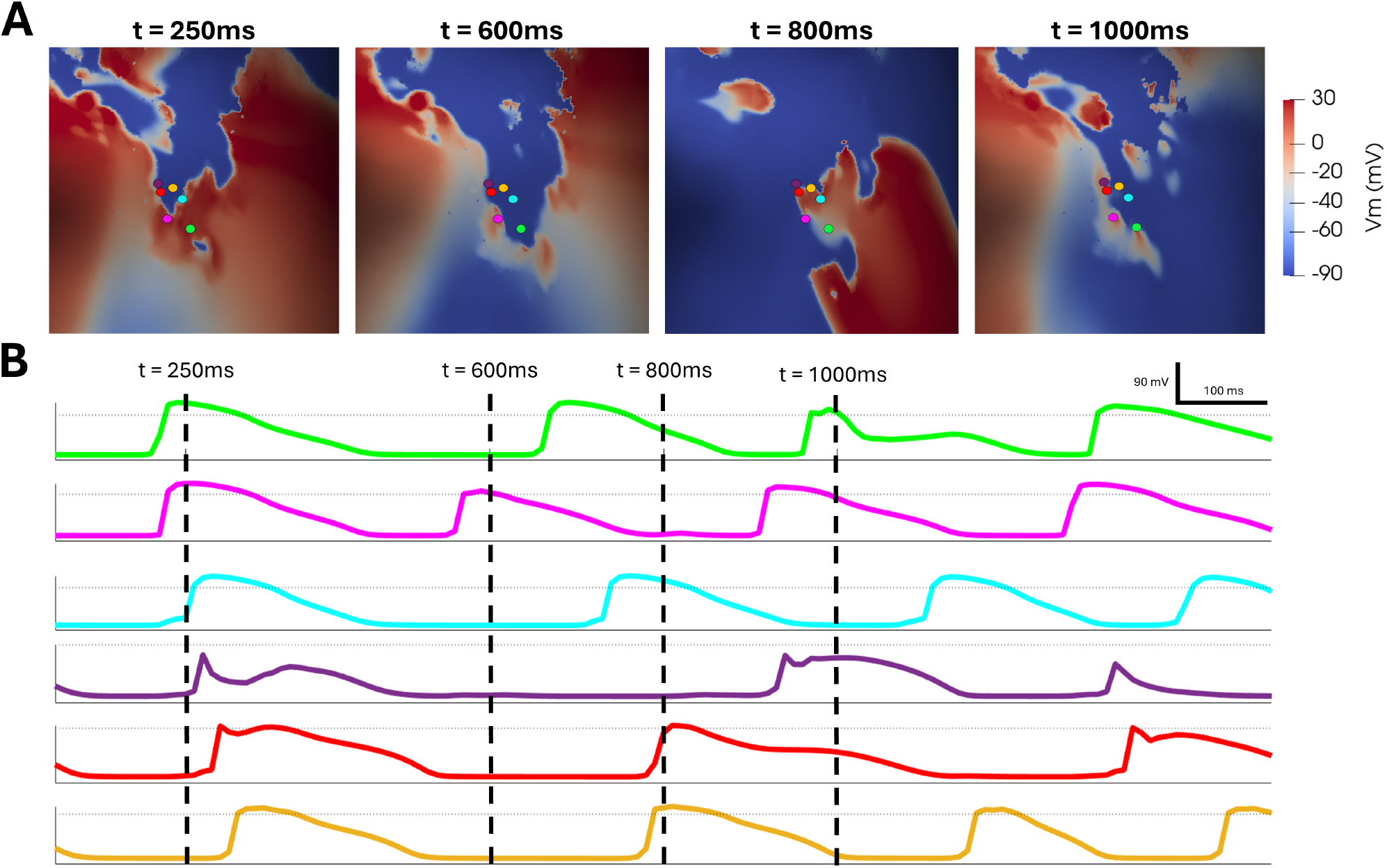
Local activation dynamics underlying RVI modulation in Patient #3. (A) Wavefront propagation at representative time points during the stimulation protocol under right stellate modulation. (B) Simulated transmembrane action potential traces extracted from the mesh nodes indicated in Panel A. Heterogeneous repolarization shortening produces spatially discordant recovery of excitability, redirection of propagation, and establishment of a functional reentrant circuit.

As illustrated in Figure 4A, following the premature stimulus, activation initially propagated through the slow-conduction BZ in a relatively organized sequence. However, under right stellate modulation, repolarization was heterogeneously shortened in myocardium near the stimulation site, resulting in spatially nonuniform recovery of excitability.

Action potential traces extracted from selected mesh nodes (Figure 4B) demonstrate that certain regions repolarized earlier and regained excitability while adjacent tissue remained refractory. Consequently, subsequent wavefronts did not reproduce the original activation pattern. Instead, transient functional block redirected propagation through partially recovered pathways, producing a progressive change in activation order across beats.

This evolving activation sequence is consistent with the establishment of a functional reentrant circuit sustained by steep local activation–repolarization gradients. Importantly, the arrhythmia did not arise from additional structural delay within the scar itself, but from stellate-induced shortening of local refractory periods in viable myocardium, thereby lowering the threshold for wavefront re-entry.

These local dynamics provide a mechanistic explanation for the marked reduction in RVI observed under right stellate modulation, illustrating how sympathetic activation reshapes functional refractoriness and promotes reentrant vulnerability independently of fixed structural substrate.

### 3.4. RVI as a marker of arrhythmic vulnerability beyond inducibility

Patient #1 provides a paradigmatic example of the added value of the RVI over inducibility-based assessment. In this patient, sustained reentry was observed across all stimulation protocols due to the presence of long, well-defined conduction channels within the BZ, rendering inducibility insensitive to additional modulatory effects, such as autonomic remodeling. Consistent with the primary hypothesis, stellate modulation produced substantial shifts in RVI_min_, RVI_1%_, and the extent of RVI< 0 regions even when inducibility outcomes remained unchanged.

As shown in Figure 5, activation of the LSG resulted in more negative RVI values across large myocardial regions. This effect is evident in the RVI maps (panel A), the leftward shift of the histogram and cumulative distribution functions, and the violin plot summarizing global RVI statistics (panel B).

**Figure 5:**
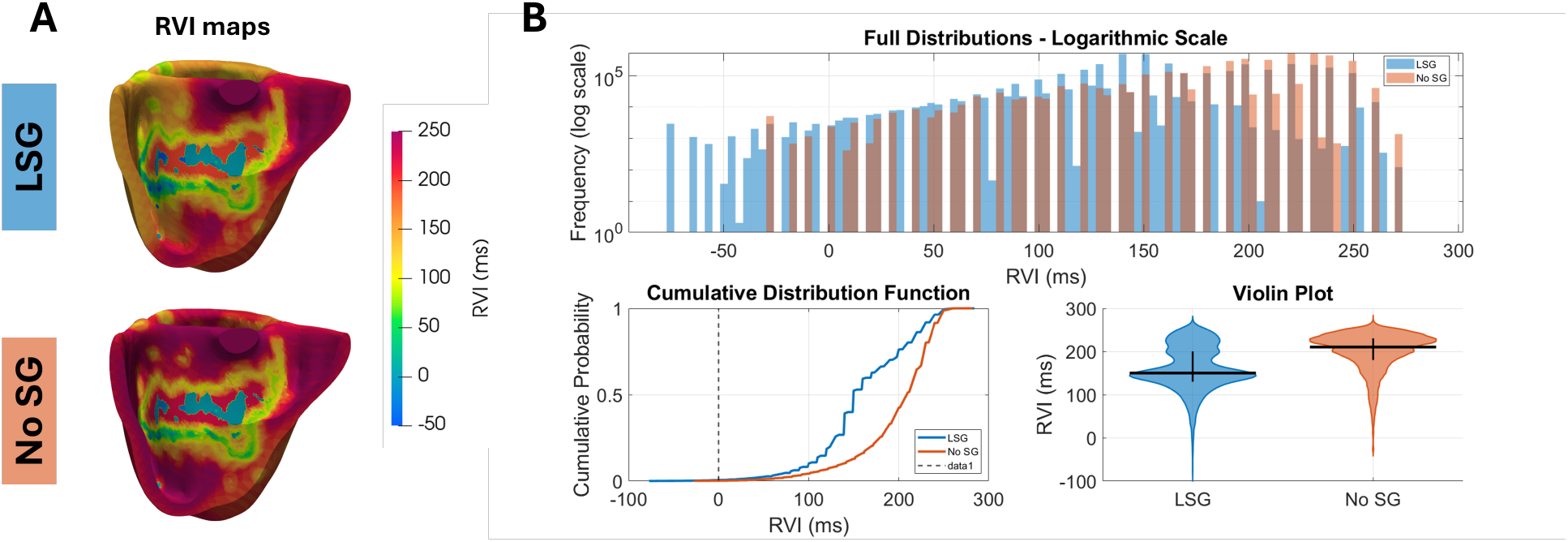
Patient #1: stellate modulation alters reentrant vulnerability despite consistent inducibility. Panel A shows RVI maps without stellate modulation and with left stellate ganglion (LSG) activation, revealing more extensive and more negative RVI regions under LSG modulation. Panel B summarizes the global RVI statistics, including histograms, cumulative distribution functions, and violin plots.

These results indicate that, even in cases where arrhythmia is always inducible, stellate modulation can unmask additional vulnerable regions that remain concealed by the stimulation protocol. Conversely, in non-inducible cases, similar RVI changes may reveal latent susceptibility not captured by inducibility alone. Together, these findings support the fact that the RVI is a more comprehensive and sensitive marker of arrhythmia vulnerability than stimulation-based inducibility.

## 4. Discussion

VAs after myocardial infarction remain a major cause of morbidity and sudden cardiac death despite contemporary revascularization and device-based therapies. In this study, we investigated how sympathetic modulation mediated by the stellate ganglia reshapes ventricular arrhythmic vulnerability in the setting of chronic post-infarction remodeling. Rather than aiming to reproduce the full spectrum of *β*-adrenergic signalling at the ionic level, our approach focused on isolating the electrophysiological consequences of APD shortening and its spatial heterogeneity within anatomically complex, patient-specific ventricular substrates. Arrhythmic vulnerability was quantified using the RVI, an electrophysiology-based metric that integrates activation–repolarization interactions and their modulation by both structural and functional factors.

The main findings can be summarized as follows: (i) stellate ganglion modulation induces pronounced, regionally heterogeneous changes in repolarization timing, leading to substantial shifts in RVI values even before the application of arrhythmia induction protocols; (ii) these changes uncover increases in arrhythmic vulnerability in both inducible and non-inducible cases, indicating that RVI captures electrophysiological alterations not fully reflected by stimulation-based inducibility alone; and (iii) the interaction between infarct location, BZ remodeling, and sympathetic modulation determines patient-specific vulnerability patterns, providing a mechanistic basis for the heterogeneous response to autonomic interventions observed clinically after myocardial infarction.

### 4.1. RVI reveals electrophysiological vulnerability beyond inducibility

Traditional in silico and clinical studies of VAs frequently rely on programmed stimulation protocols to assess inducibility as a binary endpoint. While inducibility testing remains central to identifying reentrant circuits under specific pacing conditions, it provides limited insight into the broader vulnerability landscape of structurally remodeled ventricles. In particular, arrhythmia induction may fail due to protocol constraints, limited pacing sites, or intolerance to aggressive stimulation, despite the presence of a highly vulnerable electrophysiological substrate.

Our results demonstrate that stellate ganglion modulation can substantially reshape arrhythmic vulnerability as quantified by RVI, even when inducibility outcomes remain unchanged. Patients exhibiting either consistent inducibility or complete non-inducibility still showed marked shifts in RVI distributions following sympathetic modulation, indicating that vulnerability can be dynamically altered without necessarily manifesting as inducible arrhythmia. These findings are consistent with previous computational and clinical studies showing that RVI mapping can identify regions prone to reentry without the need to induce sustained VT [16, 18–20]. Together, these results support the use of RVI as a complementary, vulnerability-oriented electrophysiological metric that captures clinically relevant information beyond inducibility alone.

### 4.2. Repolarization heterogeneity as the dominant functional driver in complex post-infarction substrates

Within the structurally complex ventricles studied here, the dominant functional effect of stellate modulation was the introduction of spatial heterogeneity in repolarization timing. Sympathetic modulation shortened APD in a regionally non-uniform manner, thereby amplifying activation–repolarization gradients, particularly at interfaces between normally conducting myocardium and regions of delayed activation within the infarct BZ. These gradients translated into low or negative RVI values, favouring wavefront–waveback interactions and increasing susceptibility to unidirectional block and reentry initiation.

This mechanism is consistent with experimental and computational evidence demonstrating the central role of repolarization heterogeneity in arrhythmogenesis [10, 21]. Importantly, recent work by [22] highlighted that *β*-adrenergic stimulation exerts multi-target effects at the cellular level, and that rotor dynamics in simplified substrates cannot be fully reproduced by isolated modifications of repolarizing potassium currents alone [22]. In contrast to such idealized settings, our patient-specific 3D ventricular models incorporate substantial structural complexity, including scar geometry, BZ remodeling, conduction velocity heterogeneity, and fibrosis. In this context, APD shortening and its spatial heterogeneity emerge as dominant modulators of vulnerability, acting upon an already intricate arrhythmogenic substrate.

Thus, rather than attempting to replicate the full ionic complexity of *β*-adrenergic signalling, our approach deliberately isolates its principal electrophysiological consequence, heterogeneous APD shortening, to examine how sympathetic modulation interacts with chronic structural remodeling at the organ scale.

### 4.3. Interaction between infarct geometry and sympathetic modulation

The impact of stellate ganglion modulation on arrhythmic vulnerability was strongly dependent on the spatial relationship between infarct geometry and regions affected by sympathetic modulation. When sympathetic effects overlapped with infarct-adjacent myocardium, repolarization shortening in tissue neighbouring slow-conduction BZ channels amplified local electrophysiological heterogeneities, resulting in highly negative RVI regions. While similar mechanisms have been described in models of acute ischemia [10], our findings extend these concepts to chronic post-infarction substrates, where fixed scar architecture and remodeled BZ regions critically shape the electrophysiological response to autonomic modulation.

In other cases, sympathetic modulation altered vulnerability through more remote mechanisms, such as reducing refractoriness near pacing sites and facilitating earlier premature stimulation, indirectly promoting reentry initiation [23]. These observations underscore that sympathetic modulation cannot be interpreted independently of infarct anatomy and highlight the importance of spatially resolved autonomic-structural interactions in post-infarction arrhythmogenesis [23, 24]. These findings are consistent with experimental evidence demonstrating heterogeneous sympathetic reinnervation and nerve sprouting in the chronic infarcted myocardium [25, 26].

### 4.4. RVI as a regional marker of vulnerability in structurally complex ventricles

A key implication of this work is the value of RVI as a spatially resolved marker of electrophysiological vulnerability that can be assessed independently of arrhythmia induction. Unlike inducibility, which yields a binary outcome dependent on protocol-specific conditions, RVI provides a continuous description of vulnerability shaped by conduction, repolarization, and tissue heterogeneity. In our cohort, stellate modulation consistently shifted RVI distributions toward more negative values, expanding the extent and severity of vulnerable regions even when inducibility did not change.

Importantly, RVI maps naturally delineate vulnerable regions rather than discrete point locations. From a translational perspective, filtering regions with RVI*<* 0 could highlight candidate areas for further evaluation within substrate-guided ablation strategies. Such regional information could be complemented by segmented conduction channels or integrated with electroanatomical mapping data [27]. Moreover, RVI analysis may also be applied post-ablation by integrating imaging data or electroanatomic mapping–based reconstructions to evaluate whether vulnerable regions persist after lesion delivery. In this context, RVI may support substrate-guided ablation strategies by identifying regions of functional vulnerability not captured by inducibility testing.

### 4.5. Implications for autonomic therapies and future modelling

The heterogeneous response to stellate ganglion modulation observed across patients provides mechanistic insight into the variable clinical efficacy of sympathetic interventions, such as stellate ganglion blockade or denervation. Clinical series have demonstrated that cardiac sympathetic denervation can reduce arrhythmic burden in patients with refractory VT, yet response rates remain heterogeneous across substrates [3, 28]. Our findings suggest that the arrhythmogenic impact of autonomic modulation depends critically on patient-specific scar geometry, BZ properties, and the spatial distribution of sympathetic effects.

Recent advances in *β*-adrenergic modeling have emphasized the importance of multi-target ionic modulation and calcium handling in less structurally complex substrates [22]. Extending such detailed formulations to patient-specific 3D ventricular models with chronic infarction represents an important future direction. Within this context, the present study provides mechanistic evidence that organ-scale sympathetic modulation interacts with fixed structural remodeling to dynamically reshape ventricular arrhythmic vulnerability.

### 4.6. Limitations

Several limitations should be acknowledged. First, sympathetic modulation was represented phenomenologically as regional APD shortening via *I*_*Ks*_ scaling rather than through full *β*-adrenergic signalling cascades. Although this approach allowed controlled interrogation of spatial repolarization heterogeneity in structurally complex ventricles, it does not capture multi-channel or calcium-handling effects associated with physiological adrenergic stimulation. Therefore, conclusions should be interpreted at the level of organ-scale electrophysiological heterogeneity rather than detailed ionic pathway dynamics.

Second, AT was defined using a fixed voltage threshold (−20 mV) for computational robustness in large-scale meshes. While consistent with prior modelling studies, alternative definitions of activation may introduce minor quantitative differences in RVI values.

Third, the study was designed to provide mechanistic insight into autonomic–substrate interactions rather than to serve as a predictive clinical validation study. While clinical VA status was included for contextual interpretation, formal evaluation of predictive performance would require dedicated prospective datasets and standardized outcome definitions.

Additionally, sympathetic territories were assigned according to standardized segmental distributions rather than patient-specific innervation imaging. While this approach ensured reproducibility, individual variability in autonomic innervation patterns may influence regional vulnerability and warrants future investigation.

Finally, the patient cohort size was limited to fourteen post-infarction cases. While sufficient to demonstrate mechanistic heterogeneity, larger multicenter cohorts will be necessary to establish generalizability and clinical utility.

## 5. Conclusion

Ventricular arrhythmogenesis emerges from the dynamic interplay between myocardial substrate and autonomic control rather than from either component in isolation. The present work underscores that sympathetic modulation, when acting on a structurally remodeled ventricle, exerts regionally distinct, substrate-dependent electrophysiological effects that cannot be inferred from anatomy or neural input alone.

The differential influence of right and LSG activity observed in this study highlights autonomic laterality as a critical, yet context-sensitive, determinant of ventricular electrical stability. In the presence of infarct-related fibrosis, sympathetic drive may amplify pre-existing electrophysiological heterogeneities, reshaping activation–repolarization patterns in ways that modulate reentrant vulnerability rather than uniformly promoting or suppressing arrhythmias.

These findings reinforce the concept that neuromodulatory therapies should be interpreted and ultimately applied through the lens of the underlying myocardial substrate. Computational modeling offers a unique avenue to disentangle these interactions in a controlled manner, providing mechanistic insight that complements experimental and clinical observations. Together, this work supports a shift toward substrate-aware, patient-specific strategies when considering autonomic interventions for VA management.

### Perspectives

#### Clinical Perspectives

Sympathetic modulation via the stellate ganglia can dynamically alter ventricular repolarization and reshape arrhythmic vulnerability in chronic post-infarction substrates. Importantly, changes in vulnerability may occur even when conventional programmed stimulation inducibility remains unchanged. Vulnerability-oriented metrics such as the RVI may therefore complement inducibility testing by identifying substrate-level electrophysiological instability. Integration of RVI-based assessment with imaging and electroanatomical mapping could improve risk stratification and inform patient selection for autonomic interventions.

#### Translational Outlook

Computational modelling frameworks incorporating patient-specific scar anatomy and regional sympathetic modulation provide a mechanistic platform to study autonomic–substrate interactions at the organ scale. Future work should focus on prospective validation of RVI-derived vulnerability metrics against clinical outcomes, integration with high-density electroanatomical mapping data, and exploration of non-invasive surrogates derived from electrocardiographic or imaging biomarkers to enable broader clinical translation.

## Data Availability

The patient-specific anatomical models used in this study were derived from previously published datasets. Due to ethical and privacy considerations associated with the underlying clinical imaging data, these datasets are not publicly available. Data supporting the findings of this study are available from the corresponding author upon reasonable request for research purposes, subject to institutional approvals and data use agreements. All data and materials necessary to reproduce the results were available to the editors and reviewers during the review process.

## Abbreviations

APD: Action potential duration
AT: Activation time
BZ: Border zone
EP: Electrophysiological
LSG: Left stellate ganglion
NoSG: No stellate ganglion modulation
RSG: Right stellate ganglion
RT: Repolarization time
RVI: Reentry vulnerability index
VA: Ventricular arrhythmia
VT: Ventricular tachycardia

## Acknowledgments

This work was supported by Grant PRE2020-091849 [MCIN/AEI/10.13039/501100011033] and “ESF Investing in your future”; Grant PID2019-104356RB-C41 [MCIN/AEI/10.13039/501100011033]; PID2022-136273OA-C33 and PID2022-140553OBC41 [MICIU/AEI/10.13039/501100011033 and by ERDF/EU]; Barcelona Supercomputing Center [IM-2021-1-0001, IM-2021-3-0001, IM-2024-1-0010 and IM-2024-2-0015]; and Teknon projects.

